# Balancing preferences and constraints around nurses shift scheduling across healthcare organisations: a qualitative study

**DOI:** 10.1101/2025.02.18.25322227

**Authors:** Hannah Ruth Barker, Peter Griffiths, Chiara Dall’ora

## Abstract

**Aim:** To understand how preferences and constraints of nursing staff, nurse managers and hospital directors interact and influence decisions around nurse shift patterns

**Background:** Globally, many nursing staff have expressed intention to leave the profession and high levels of stress and burnout. Shift patterns are often cited as a key driver of negative nurse outcomes. How preferences and constraints of staff groups involved in nurse shift scheduling interact to shape decisions remains unexplored.

**Design:** Qualitative study. We interviewed 17 nursing staff, 5 nurse managers and 6 hospital directors.

**Methods:** Staff were recruited through 5 NHS Trusts based in the North, Midlands, East and South of England. The results were analysed using inductive and deductive framework analysis.

**Results:** Three themes were identified that influenced nurses’ decisions for shift scheduling: 1/ Balancing Choice with Consistency, Predictability and Flexibility; 2/Adequate Rest and Recovery Between Shifts; and 3/Enjoyment and Engagement at Work. There was a meta-theme of conflicting priorities for shift scheduling between nursing staff, nurse managers and hospital directors across these three themes.

**Conclusions:** Different priorities for shift scheduling of nursing staff, nurse managers and hospital directors created potential or actual conflict. Solutions appeared to consist of compromises where priorities of different staff groups were simultaneously upheld to an acceptable level. This occurred through collaborative interactions across staff groups.

**Implications for Practice:** Nursing staff, nurse managers and hospital directors can facilitate choice and flexibility and navigate conflicts in nurse shift scheduling through collaborative approaches that enhance awareness and effective communication.

**Reporting Method:** Qualitative data was reported using the EQUATOR checklist COREQ. No Patient or Public Contribution

What does this paper contribute to the wider global clinical community?

Collaborative approaches can influence conflicting priorities of staff groups in relation to nurse shift scheduling.

Staff understanding other staff priorities for nurse shift scheduling enhances awareness.

## Introduction

Internationally, there is evidence that the nursing workforce is facing increasing burnout, stress and absence, alongside fundamental concerns about unsafe staffing and poor working conditions (Buchan & Catton, 2023). This is problematic because a depleted nursing workforce presents a major challenge to the delivery of high-quality services, now and into the future. In England, a significant proportion of nursing staff have declared plans to leave their jobs and/or the profession (NHS England Digital, 2024). A lack of work life balance is a key reason for staff leaving the nursing profession (England Digital, 2024) and links between working unfavourable shift patterns and reports of poor work-life balance have been highlighted (Emmanuel et al., 2023; Karhula et al., 2018). The challenge for NHS managers is to meet cost and efficiency targets through effective rostering while also addressing staff work-life balance. This is important because, in recent years, prioritising cost and efficiency targets alone has negatively affected nursing staff morale and increased sickness rates (The King’s Fund, 2023).

## Background

Previous studies, not specific to nursing, have linked staff choice in shift scheduling to wellbeing and work life balance (Nijp et al., 2012). Flexible work arrangements that include more personal choice have been associated with better physical health and reduced absenteeism (Shifrin & Michel, 2022). Choice in shift scheduling has generally led to positive outcomes for nursing staff, including reduced sickness absence, burnout, turnover intention, and work-life conflict (Dall’ora et al., 2023; Grøtting & Øvergård, 2023; Karhula et al., 2018; Kida & Takemura, 2024; Turunen et al., 2020). However, some research has reported uncertainty on whether increased choice in shift scheduling consistently improves nursing staff wellbeing (Karhula et al., 2020; Wynendaele et al., 2021).

In addition, there is limited evidence around how elements that drive choice and preference around shift patterns for nurses interact with those of nurse managers and hospital directors in the organisation to shape decisions. A large qualitative study by Emmanuel et al. (2023) found that nurses’ shift pattern preferences were frequently related to their priorities outside of work, consistent and predictable shift patterns, and flexibility over working time. Similarly, a recent qualitative study highlighted that nurses favour shift patterns that align with their personal lives but warned these could be prioritised over fatigue management recommendations (Booker et al., 2024). To understand how preferences and constraints between nurses, their managers and hospital directors interact we will consider conflicts, compatibilities and compromises made between different staff groups in the organisation. When we describe conflict in this paper, we use two terms, potential conflict and actual conflict. By potential conflict we mean conflicting needs or priorities, where different preferences discord creating situations that are ready for conflict. When we state actual conflict, we refer to when the conflicting priorities or choices are expressed as an open disagreement, involving frustration or tension. Importantly, potential conflict or conflicting priorities do not mean conflict is inevitable. These definitions are aligned to seminal work on the nature of conflict (International Sociological Association, 1957).

Therefore, the aim of our work is to understand how preferences and constraints of nurses, nurse managers and hospital directors interact and influence decision making around shift patterns.

## Methods

This was a qualitative study and the EQUATOR checklist Consolidated Criteria for Reporting Qualitative research (COREQ) was applied (Tong et al., 2007). A purposive stratified sampling framework was used to ensure a variety of settings by geography and healthcare speciality. The study advert was shared on the National Institute for Health and Care (NIHR) Clinical Research Network, and Research and Development (R&D) offices contacted the research team and self-selected to the study. We recruited five diverse Trusts based in the North, Midlands, East and South of England. Two had specialities in Mental Health, one in Paediatrics and two were general NHS Foundation Trusts.

In each Trust, the R&D team or local Principal Investigator (PI) advertised the study through tailored recruitment channels. Eligible participants were nursing staff (registered nurses, nursing associates and healthcare support workers) in participating Trusts who worked shifts in inpatient settings; ward managers who worked in inpatient settings and hospital directors who worked in HR or Finance settings. Ethical approval was granted by the Health Research Authority (IRAS ID: 327884) and from the University of Southampton ethics committee (ID: 81204), all participants self-selected to the study, and gave online or audio consent. Data was collected between October and December 2023.

Seventeen nursing staff working shifts (including registered nurses and health care support workers), five nurse managers in charge of wards or departments, and six hospital directors were interviewed. All interview were semi-structured and lasted between twenty to fifty-five minutes. Nursing staff and nurse managers were interviewed individually and were asked to share their experiences and perceptions of nurse shift patterns: what a typical shift pattern looked like, what was most satisfying and most frustrating or challenging about the shift pattern, what choices and factors made an ideal shift pattern, what were non-negotiables in shift patterns and to describe well-being and work-life balance. Hospital directors conducted one group interview (lasting 55 minutes) with similar questions. A group interview was conducted rather than a focus group to support consistency in the data collection and analysis.

The interviews were conducted by researcher (HRB), who had previous experience in qualitative data collection & analysis, via Microsoft Teams video call and recorded. Qualitative interviewing continued until there was data saturation in nursing staffs’ responses. The interviews were transcribed verbatim (HRB), read and re-read for familiarity and the results were analysed using inductive and deductive framework analysis (HRB, CDO and PG) (Ward et al., 2013). The ontological position taken, compatible with these methods, was subtle realism. This offered a balanced view on the data, maintaining that the social world existed independently of an individual’s perspective, meanwhile recognising that an individual’s understanding of the world was shaped by their unique perspectives, experiences and interpretations (Hammersley & Atkinson, 1995). Through qualitative research, understandings were made accessible as the researchers interpreted the participants’ explanations (Ward et al., 2013). No relationships with participants were established prior to the interview and participants knew only that the researcher had an interest in the research topic. HRB used techniques of reflective journaling throughout data collection and analysis to develop the code creation and provide awareness of her potential bias to enhance rigour, HRB also observed her own characteristics that could affect the qualitative analysis. Other researchers (CDO and PG) were consulted to discuss the development of the codes and themes, for example initially the themes were organised deductively: i.e. shift patterns, rostering, ancillary practice, pay, wellbeing and life outside of work; however, on further discussion these themes and ensuing sub-themes were shifted and condensed inductively into three overarching themes and one meta theme. The audit trail that arrived from the framework analysis method, alongside teamwork and collaborative analysis, enhanced dependability and confirmability (Tobin & Begley, 2004). Finally, nurse leaders across the five Trusts engaged with the study’s findings, confirming the results resonated with their experiences and perspectives.

## Findings

We identified three overarching themes influencing nursing decisions regarding the deployment of shift patterns in hospitals: Balancing Choice with Consistency, Predictability & Flexibility; Adequate Rest and Recovery between Shifts; and Enjoyment and Engagement at Work. There was also a meta-theme of conflict across these three themes: Different Priorities & Constraints for Shift Scheduling of Nursing Staff, Nurse Managers and Hospital Directors Created Potential or Actual Conflict. The meta-theme showed potential and actual conflict endured due to different priorities and constraints of staff groups across the organisation, and that where compromises were actioned potential conflict did not become actual conflict. Different interactions between staff groups to potential conflict affected whether or not actual conflict was realised. To highlight the meta-theme of conflict within the three overarching themes in the results we use bold font. We reported the accounts from three different staff groups as follows: nursing staff (ID), nurse managers (NM), and hospital directors (D).

### Theme 1: Balancing Choice with Consistency, Predictability and Flexibility

Nursing staff valued consistency, predictability and flexibility in their shift patterns. Different **priorities** and **constraints** among nursing staff, nurse managers and directors affected how these attributes were balanced in nurse shift scheduling choices.

Nurses valued a consistent shift pattern. Nursing staff emphasized the importance of avoiding ‘random’ shifts. **Actual conflict** expressed as frustration or tension was evident when schedules included “*sporadic*” (ID8; ID10) or “*random*” (ID9) shifts, particularly when days and nights were mixed, as this was disorientating for staff and meant they had difficulty planning their lives outside of work. Generally, nurses expressed needing routine in their lives for planning and quality of life outside of work: “*It’s all shuffled and there’s no routine in your life*…*When you work when you have a routine in your life*… *I go out to my friends*… *I go to gym*…*I reboot*” (ID7). Having a consistent schedule made planning outside of work easier for nurses.

A consistent schedule could look different depending on nurses’ individual choice. For example, consistency might mean always having Tuesday and Friday evenings off for childcare purposes, or it could involve working 90% night shifts. For nursing staff, non-negotiable **priorities** for shift scheduling often related to commitments outside of work, such as caregiving responsibilities, income needs, and health and wellbeing: *“I actually was doing most night shifts when I returned to work after maternity leave because of my husband was doing day shifts, was working nine to five. So I had to do night shifts just to balance who takes care of our baby*.*”* (ID30).

Nonetheless, **conflicts** arose because managers could not always accommodate each individual’s version of a consistent schedule: “*They lost about 3 qualified nurses to a different trust because they weren’t given…set days for…childcare*” (NM14)). This necessitated a degree of **compromise** from both the nurses and the organisation. For example, while individuals needed to be willing to work weekends, the organisation could provide a level of consistency regarding weekend work: *“I like the fact that our trust has a set rule that we should get at least two weekends off every month”* (ID23).

Nurses valued predicting their work schedules well in advance. An aspect that was **compatible** across participant groups was planning and releasing rosters in advance. It worked for staff and managers: “*The earlier that you can actually get them approved, signed off, any vacant shifts uploaded to NHSP and filled with bank rather than having to go out to agency, is a lot better*” (D4). For the organisation, this was better for continuity of care and finance. For staff, when rosters were approved in advance it was useful for planning their lives outside of work which also contributed to their wellbeing: “*if you know what days you’re working and which days you’re off, then it’s easier for you to plan*” (ID2). Regarding rota lead time, the value of “fairness” was shared between nurses, their managers and the wider organisation, and unfairness in rostering was linked to rostering at short notice: “*we have a lot of unfairness around rostering at those periods of time (i*.*e*., *Christmas, holidays) because they (i*.*e. the rosters) are done at such short notice* (D3).” For directors and nurses, fairness was not an abstract value. Directors and nurse managers conceived fairness as a “win-win” that would enable an efficiently run organisation: “*people can plan their lives further in advance so that we can make sure that people are fairly managed and we’re organised* (D3)”. Meanwhile, nurses acknowledged that fairness in the roster made not always having their preference acceptable: “*If I could choose not to work weekends, I probably would. But then that means someone else would have to work them, and that’s not fair*” (ID8). In essence, a predictable schedule released well in advance with fair scheduling offered some protection to offset **actual conflict** from shifts that were not desirable: “*I think longer is better… if you do need to shift swap or something you have a bit of time to play with*” (ID6).

Nurses valued flexibility from their managers. For nurse managers one key priority was balancing staff preferences on the off duty for a happier workforce: *“If I make sure everyone’s kind of like happy with the off duty that anxieties aren’t attached to that. Everyone’s fairly happy then you’ve half the battle”* (NM14). The goal wasn’t for complete perfection but that ‘everyone’s fairly happy’; this indicated the ability for nurse managers to be flexible and to balance the expectations and preferences of their staff, and the ability of nursing staff to **compromise** and to be flexible with their ideal shift pattern when enough of their individual preferences were upheld. When nurse managers attempted to accommodate the preferences of their nursing staff, they engaged in one-to-one communication to understand and raise awareness of their staffs’ preferences: *“it’s about assessing the individual. I don’t think there is a blanket rule”* (NM3).

Additionally, nurse managers found that supporting staff preferences could improve staff retention. Some managers explained that, based on their experience, organising shift patterns to acknowledge individual preferences reduced staff sickness and improved retention. In this instance, reducing staff sickness was a key driver for arranging shifts according to staff needs: *“Once you get to know your workforce*…*you can play what they want and then they’re going to attend* …*my sickness went from double figures percentage to like 2*.*3%”* (NM14). Similarly, when flexible working arrangements (a routine of consistent shifts agreed with a line manager who was flexible to the needs of the individual) were in place, they improved satisfaction in work and work life balance for nurses: “*I do have agreed flexible working. Suits me and it suits my needs. I’m able to go to the gym on a Monday night* … *I’m able to see family more*…*I’m able to, you know, go on holiday*” (ID21).

Other factors contributed to how much flexibility nurse managers were able to offer their nursing staff. Nurse managers faced **constraints** in changing shift patterns due to other staff requests, staff skill mix, establishment targets, staff sickness, and vacancy levels: e.g. *‘when we do the off duty, we have to make sure the ward is covered first and* …*only then, we can honour those hours like you know the additional hours for the learning opportunity’* (NM11). The hospital directors also recognised **constraints** due to the wider health service demands for Trusts to grant their staff more flexibility: “*Now that the NHS are really pushing flexible working it’s really quite difficult to do that (…) Especially one of our significant challenges is term-time only contracts, which is what we offer, because it’s very difficult to accommodate that on a ward because predominantly there’s, you know, lots of females that work on our wards who are you know, provide the main care to children. Because if you have maybe nine nurses on one ward it’s difficult to grant them all term time only contracts, so it’s about finding that balance*” (D2). Nursing staff did understand some of these **constraints** due to communication with their managers “*They are quite approachable due to myself having children. But It’s hard to change because of requests from other members of staff and sickness as well*.” (ID29). Despite these **constraints**, the level of flexibility that managers were willing to prioritise for their staff varied: “*the band six. She does try to give me the Thursday off if I’m working nights or the Friday morning off if I’m working days. But the band seven does not*” (ID29).

**Actual conflict** existed when nursing staff felt let down by their managers’ complete rigidity in responding to their specific needs: “*Doing 2 long days were too difficult for me in a row because I was hardly sleeping* …*So I took this issue to my superiors and I asked them if I could if I’m working Monday a long day, can I have a day off on Tuesday and work again on Wednesday so that, you know, it’s not too tiring for me. But it wasn’t, it was not possible*… *I’m asking for just a gap between the days and they said that’s not possible*” (ID7). When nursing staff felt ignored by their managers: *“it’s not our issue, swap your shift”* (ID8) stress and **frustration** arose.

Key **priorities** for hospital directors centered on safety targets (i.e. safe staffing levels and incident reports) and budget: patient safety was “*the top priority*” (M4) and then to save “*money and make services more efficient*” (M4). When hospital directors considered the different **priorities** for shift scheduling of nursing staff, and applied measures to ‘soften’ **potential conflict** to prevent it becoming **actual conflict**, these were often perceived by directors as being at odds with service provision goals and viewed as a last resort. Due to high levels of nurse turnover, directors were willing to **compromise** on Trust policies that had been originally implemented to ensure patient safety: “*We didn’t have night shifts only contracts, it was not allowed, but now we have softened because we’ve realized, you know, we’ve gotta go with what individuals want rather than it to be what the organisation wants. We’re in a buyers’ market and that is what’s pushing our behaviours, we are purely organising our services around the individual rather than necessarily what the service or best practice might tell us. And that’s because we’re in a corner. So that’s I think what’s driving us”* (D3) and “*So people will move and go to another organisation because they will accommodate the longer shifts or the flexible working etcetera. So it’s almost like it’s for the members of staff. It’s a buyers’ market for them at the moment*” (D1).

In summary of Theme 1: nursing staff wanted choice in their shift scheduling for a consistent shift pattern that suited their individual needs. Individual needs were understood through effective communication and raised awareness between nursing staff and their managers. However, not all individual preferences could always be met due to organisational **constraints**. To balance this, nursing staff needed some flexibility to work undesirable shifts, and nurse managers needed flexibility to provide as much consistency for desirable shifts as was possible meanwhile balancing expectations in relation to organisational **constraints** and offering predictability to offset **compromise** on undesirable shifts.

### Theme 2: Adequate rest and recovery between shifts

A key theme for nursing staff was achieving sufficient rest and recovery between shifts. The challenges of working as a nurse were universally recognised by all parties, as the job was tiring emotionally, psychologically and physically. If other **priorities** of nurse managers and hospital directors resulted in rotas that prevented nursing staff from getting adequate rest between shifts, this led to **actual conflict**. For example, if shift finish times were too late in the evening to unwind properly for sleep before a sequential shift; or when there was insufficient rest after a block of shifts, for example after two long days. What constituted adequate rest time between shifts differed from person to person, but what was evident for everyone was that it needed to be enough, where the time for recovery fell short the effect on following shifts culminated in fatigue and stress. When too many shifts were ‘bundled’ together over consecutive days nurses felt drained of energy “*that would mean that you have something crazy like 6 shifts together without a break*” (ID9).

What nursing staff found most satisfying about their shift patterns included their time off “*the days off, I think by doing long days it means I get more days off so I find that better for a work life balance”* (ID17). Recovery between shifts was key to achieving work life balance and this included wanting to sleep well and rest sufficiently, to emotionally recuperate, to get organised outside of work (i.e. to provide for and be present with their families) and to socially recuperate (i.e. to enjoy weekends with friends and family). Work-life balance meant something different for everyone, but it was clear that shift patterns significantly affected it. When work-life balance was deemed successful, it positively impacted overall wellbeing: “*My wellbeing, I would say. Is good when my shifts are within a sensible sort of pattern during the week*… *when I have weeks* …*when I had both weekends either side of three shifts, then a training day, then that’s when my wellbeing sort of takes a bit of a toll and I feel like I don’t have enough time to recover properly. So, I go into the next week being really tired*” (ID9). Some nursing staff resorted to flexible working arrangements to achieve a better work life balance. Nonetheless, there were cases where individual **constraints**, particularly childcare responsibilities outside of work, made achieving work-life balance elusive, even with a flexible working arrangement: “*So I do 3 long shifts a week, … it tends to be 2 long days in the week and then again because of childcare commitments and things. It’s always a shift at the weekend, whether that’s a day or a night…. It’s not working for me at all at the minute. Just work life balance is horrible basically*” (ID6).

Throughout the nursing staff interviews, a theme of earlier long day finishing times was identified. Specifically, the shift finish time appeared to significantly impact staff work-life balance. Finishing earlier at 7.30pm or 8pm offered staff a chance to unwind and rest before bedtime: “*Your adrenaline is going and then you get to like 7:00pm* … *and you’re just like mentally drained. But then you still have two hours more to go, so it feels like it just goes on and on some days*” (ID13) and “the times where you know if it actually finishes at half seven. And you go home… You’ll get a couple of hours in the evening before you have to get up the next day” (ID21). The solution to this **conflict** involved finding **compatibility** between the nursing staff’s priority of completing their long shifts earlier, the nurse managers’ priority of maintaining a satisfied workforce, and the organisation’s priority of cost savings: “*the end time of a shift pattern is a big difference in how people feel… my argument to get that time and pushed back is actually we save on budget*” (NM1).

### Theme 3: Enjoyment and Engagement at Work

Other shift pattern **priorities** for nursing staff centered around being able to spend meaningful time with patients and the wider interdisciplinary team: “*interacting with the patient more*…*spending more time with them and learning more about them* …*And actually seeing*…*people*… *my manager*…*like doctors* …*just knowing how the ward works* … *it’s much, much better*” (ID18). Interestingly, nurses with shorter daytime shift patterns (such as early and late shifts or management hours) expressed less often experiencing shortfalls in **priorities** relating to patient interaction and team working compared to those working long day or night shifts. Any shortfall in the team, handover, or professional expertise was felt more acutely with long day shift patterns or night shifts, with nurse managers highlighting that creating a positive team climate was crucial for an enjoyable shift: *“everybody knows* …*we’re gonna be here for this long together. And we only have three people on shift which I think kind of reinforces that you need to be a lot more of a tighter team because then if one person’s upset or one person disrupts something that can cause quite a big ripple effect”* (NM32).

Some nurse managers expressed the importance of giving staff ‘time off the floor’ in the roster despite it not being ‘normal’ practice: “*it makes them feel appreciated about you know what they’re doing because it’s not normal for a band 5 staff nurse to have a management day, and I think, well, they need it though. They need time off the floor to be able to do what I want them to do*” (NM14). Other ways that nurse managers offered support successfully was through conversations whereby they listened, understood and acknowledged the needs of their staff and were able to suggest a new direction, a temporary change or a training opportunity: “*have you thought about this course or that training opportunity*” (NM32).

Opportunities offered by Trusts for wellbeing were perceived as inaccessible to nursing shift workers who required it most: “*health and wellbeing department and they put on like lovely events*…*and they forget about shift workers*… *I can’t leave the unit for an hour and go on a lovely midday walk with you. You know, we’ve got a staff gym just opened. It’s open four till seven. I don’t finish my shift until half seven*” (ID6).

## Discussion

Our study was the first to investigate how different priorities of nursing staff, nurse managers and hospital directors interact and shape nurse shift patterns, revealing potential conflicts, actual conflicts and occasional compatibilities. Despite interviewing a variety of nursing staff and nurse managers with different backgrounds and lifestyles, we developed three overarching themes that influence nursing staffs’ decision making in shift scheduling: Balancing Choice with Consistency, Flexibility, and Predictability; Adequate Rest and Recovery between Shifts and; Enjoyment and Engagement at Work. Novel to our study is a unique meta-theme relating to conflicting priorities, between staff groups (nursing staff, nurse managers and hospital directors), across the three overarching themes. We showed potential and actual conflict existed due to different priorities and constraints of staff groups (nursing staff, nurse managers and hospital directors) in the organisation, and that where compromises were possible potential conflict did not become actual conflict. For example, nurses’ preferences could be reasonably met, through collaborative approaches to compromise, to support the organisations’ priority of retaining staff. However, in some cases, when no compromises were made, nurses left their jobs.

## Interpretation of the findings

### Balancing Choice

Recent qualitative work has highlighted features that are valued by nurses in shift scheduling such as rota consistency, work time control and flexibility (Emmanuel et al., 2023), and having their preferences acknowledged (Ejebu et al., 2021). What is significant about our research is that although we captured these elements similarly, we further interpret how these different, but overlapping principles, worked together. In the theme Balancing Choice, the three elements of consistency, flexibility and predictability worked together to create an approach to choice.

Consistency provided a starting point of setting clear expectations and predictable outcomes. Flexibility from nursing staff and managers allowed for adaptation and responsiveness to less desirable shift patterns and staff individual needs respectively. This indicated that to balance choice with consistency and flexibility expectations could be managed through effective communication and raised awareness between staff groups (nursing staff, nurse managers and hospital directors). Nursing staff could expect some consistency in their shift patterns for their individual needs, but also that some give-and-take would be required to deliver within organisational constraints.

Predictability surfaced from the interplay of consistency and flexibility as an approach that nurse managers could leverage to make less desirable shifts more acceptable to nursing staff, by managing expectations effectively through reducing uncertainty and providing some stability, but also through ensuring fair processes for all nursing staff team members.

Our research is also significant because it expands the narrative around choice in shift scheduling and wellbeing in nurses. Our study indicated choice in nurse shift scheduling was consistently perceived by nursing staff as a benefit to wellbeing and work life balance, it indicated people may feel better when they believe they have more choice. These results sit uniformly with most other research exploring nursing staff choice in shift scheduling and wellbeing outcomes (i.e. more job satisfaction, less sickness absence, more work-life balance, less fatigue, less burnout etc.) demonstrating positive associations between choice for shift scheduling and wellbeing outcomes (Dall’ora et al., 2023; Fond et al., 2023; Karhula, Hakola, et al., 2018; Lamont et al., 2017; Turunen et al., 2020). However, in our study, external constraints, such as caring responsibilities, sometimes meant that despite choice being offered at work, nurses remained stuck in relation to work life balance. These findings add novel insight and may explain why some results on choice for nurses in shift scheduling and perceived wellbeing showed little effect (Karhula et al., 2020). Karhula et al. (2020) did not adjust for lifestyle constraints outside of work on participatory scheduling in comparison to traditional scheduling thus our work is useful to highlight possible reasons for these results in the absence of studies which address this evidence gap and that can be more widely applied to healthcare settings.

Research on flexible working arrangements has shown that managers support the concept due to the perceived benefits to their individual staff, (such as: trust, confidence, increased morale and mutual respect) (Mercer et al., 2014). Flexible working arrangements have been touted as the solution to retention challenges (NHS, 2020), but with little evidence to guide implementation in complex health care systems. Our research indicated issues were present, i.e. staff preferences could not be perfectly granted with flexibility due to organisational constraints. Other research concurs that in shift work and nursing settings the physical and immediate nature of the job (24-hour face to face care) was a barrier to flexible working arrangements compared with 9 to 5 office-based settings (Mercer et al., 2014). The current study is instrumental in understanding what shift pattern modifications might look like. Through expanding understanding of how staff groups across the NHS interact in relation to shift scheduling it is clear there is a role that consistency, flexibility, predictability, fair processes, effective communication and raised awareness play for all staff groups, despite conflicting priorities, in making choices and compromises related to shift scheduling acceptable.

### Rest & Recovery between Shifts and Enjoyment & Engagement at Work

Our results showed the importance of rest and recovery between shifts and enjoyment and engagement at work for nurses. For example, actual conflict occurred when there was not enough rest for nursing staff between shifts, and too many shifts were bundled together. Additionally, many nurses felt that long days facilitated this rest, providing enough down time away from the ward. These themes highlight a shift away from focusing solely on pay or days off and indicate that nurses are increasingly prioritising their wellbeing and that their job satisfaction was also important to them. Perhaps these themes indicate that nurses recognise the impact that wellbeing has on their ability to provide quality care in a sustainable way, which in turn offers job satisfaction. Certainly, our research shows nurses have an interest in schedules that support their physical and mental health, and that this supports them to be fulfilled in their work life balance. However, similar to other research (Emmanuel et al., 2020), our research also indicated shortfall in positive team dynamics, handover and training opportunities with long day shift patterns. Our study uniquely showed that although these elements were still valued by nurses, frequently nurses would nonetheless prioritise maintaining a long day shift pattern for the benefit of work-life balance. Our study indicated that when nurse managers gave their staff ‘time off the floor’ for training opportunities it supported their staff to deliver quality care when they returned to their shift work. Our research shows that furthering understanding of how to strengthen team dynamics, training, communication and handover in a ward that employs long shifts is an imperative multi-pronged approach to shift scheduling and retaining staff.

Additionally, in our research, some nurses were constrained by other priorities outside of work, for example income requirements, or caring responsibilities. A recent scoping review highlighted similar findings, and suggested these constraints could lead to sacrificing job satisfaction to balance demands (Ejebu et al., 2021). One qualitative study expressed concern that allowing nurses complete autonomy in choosing their preferred shifts could lead to prioritising other needs over their individual well-being (Booker et al., 2024). In contrast to Booker et al. (2024), our study, in the theme Rest and Recovery between Shifts, showed how important rest and recovery was to nurses for their work life and their life outside of work; additionally, the theme Enjoyment and Engagement at Work, although to a lesser extent, indicated that nurses had dependable priorities related to their engagement at work. This offered a different perspective on nursing staff who are given choice for their shift patterns, and does not support the idea that their wellbeing would be impaired because of it.

### Meta-theme: Conflict

Within the three overarching themes we were able to see examples of when potential for conflict remained as such, with successful compromises from different staff groups, and when it became actual conflict. The most obvious trend was when there was no flexibility at all, when there was complete rigidity on the priorities of different staff groups actual conflict resulted. For example, if only rostering targets were considered in shift scheduling decisions, or if an individual with a non-negotiable need was met with no flexibility. Sometimes a small tweak was enough, for example an earlier shift finish time or a specific shift pattern, i.e. no consecutive long day shift patterns. However, it was also clear that getting small changes agreed in the organisation could prove difficult. In the current research, we were not able to delve deeper into these barriers to change, but this would be a worthwhile direction for the future.

When there were compatibilities between nursing staff and the organisation, these were nonetheless underpinned by different priorities. For example, managers embraced aspects of flexibility while acknowledging challenges, but tended to frame this in terms of the benefits to operational factors, such as reducing turnover, rather than focussing on direct effects for staff wellbeing. These different priorities and constraints perpetuated potential conflicts across the organisation and finding compromises was necessary to avoid actual conflict to continue delivering hospital services and to avoid exceedingly high and unmanageable nursing staff turnover challenges. Other studies have qualitatively explored organisational constraints for nurse shift scheduling to inform rostering models and algorithms (Ang et al., 2018; Klyve et al., 2023; Leung et al., 2022; Mischek & Musliu, 2019), and their results were similar to the constraints identified in our study. One case study concluded hospitals needed more flexibility to navigate constraints, they suggested creating room to manoeuvre by increasing the number of staff and relaxing certain rules (such as allowing consecutive shifts in certain situations) (Aeschbacher et al., 2023). Our research similarly showed examples of the role of compromise in softening potential conflicts. Nurse managers indicated examples of improved staff retention and reduced sickness absence when balancing nurses’ preferred shift patterns alongside the staffing requirements for the ward. Meanwhile, nurses offered examples of leaving jobs when flexibility and balancing of the roster didn’t happen. Our study showed that each staff group’s understanding and upholding of the other staff groups’ priorities to an acceptable level helped maintain the different priorities as potential, rather than overt, conflicts. Effective communication between staff groups that raised awareness, and created adjustments in expectations and fairness facilitated this flexibility.

## Implications of the research

In the current study, we offered insight into how different elements in shift scheduling choices connected. However, this work was qualitative and therefore not generalizable. Rigorous quantitative research is needed to understand more objectively how different aspects in choices for shift scheduling are prioritised by nursing staff. For example, understanding how nursing staff trade off the importance of a consistent schedule, a predictable schedule offered in advance and their shift length (e.g. 12-hour shifts), to uncover whether patterns exist to advance shift scheduling guidance.

Our research also highlighted the under researched area of constraints for nursing staff outside of work (frequently these constraints were child or adult caring responsibilities), that affected work life balance and shift scheduling choices. Future research should aim to adjust for these variables, when measuring wellbeing and work-life balance outcomes in relation to choice in shift scheduling.

Work life balance and wellbeing were valued by nurses in the current study and were particularly highlighted in the rest and recovery between shifts theme, as well and the enjoyment and engagement at work theme. These attitudes may reflect a broader societal shift towards prioritising well-being and work-life balance (Shifrin & Michel, 2022). Hospitals need to be ready to respond to these changing priorities, and innovative scheduling processes based on up-to-date research can support this shift. For example, if nurses become more vocal in relation to their shift scheduling needs, power dynamics could change between employees and employers prompting more collaborative scheduling processes. Given the hierarchical and arcane power structures still maintained within the NHS, this may well be a catalyst for change and lead to untold benefits. Furthermore, research on how priorities for work life balance and wellbeing differ between different age groups, experience levels and nursing specialities would be worthwhile to understand patterns in attitudes, to better address them.

Further research to develop interventions to test the findings of the current study relating to compromise and conflicting priorities of different staff groups for shift scheduling in the NHS is worthwhile given the need for evidence-based shift scheduling guidance. Following the development of evidence-based interventions, robust cluster randomised control trial designs would ideally evaluate their effectiveness.

## Limitations

Individuals may not always be fully open in their responses as the influence of speaking with an unfamiliar person and knowing that their experiences are being recorded may moderate their responses (Burr, 1995). However, the researcher used her previous experience in interviewing, to encourage the participants to feel relaxed and safe in their environment to support authentic accounts. Additionally, rigour was supported through reflective processes (i.e. reflective journaling) and dependability (i.e. discussing results with other researchers) where possible during the data collection and analysis to enhance the trustworthiness of the results.

Furthermore, we don’t know anything about actual behaviours and choices. The interviews provided us with perceptions of what was important to nurses in relation to shift scheduling and how these aspects interacted with other staff groups’ shift scheduling priorities. However, we do not have research showing the actual behaviours and choices that nurses, nurse managers and hospital directors make for shift scheduling, and these may differ from the perceived behaviours and choices.

## Conclusions

Globally health care systems are losing nursing staff due to poor work-life balance. Our research points towards the importance of choice for nursing staff in shift scheduling to promote wellbeing and work-life balance. The study shows that shift scheduling is a contentious issue and is a point of potential and actual conflict between the organisation and nursing staff due to different priorities and constraints. Compromise provided a solution to reconcile actual conflict, through upholding priorities of different staff groups to an acceptable level. Compromise was usually achieved through effective communication and collaboration between staff groups. This led to greater awareness of the priorities of different staff groups, and created adjustments in expectations and fairness, which facilitated flexibility across the organisation. To support effective shift scheduling evidence-based guidance is needed to navigate complex healthcare settings with constraints unique to other sectors.

## Data Availability

All data produced in the present work are contained in the manuscript

